# Examining the relationship between adolescent health behaviors, brain health, and academic achievement using fNIRS

**DOI:** 10.1101/2020.09.08.20190835

**Authors:** Mia Papasideris, Adrian Safati, Hasan Ayaz, Plinio Morita, Peter Hall

## Abstract

**Background:** Several adolescent health behaviors have been hypothesized to improve academic performance via their beneficial impact on cognitive control and functional aspects of the prefrontal cortex (PFC). Specifically, exercise, restorative sleep, and proper diet are thought to improve PFC function, while substance abuse is thought to reduce it. Few studies have examined the relationships among all of these in the same sample, while quantifying downstream impacts on academic performance.

**Objective:** The primary objective of this study is to examine the association between lifestyle behaviors and academic performance in a sample of adolescents, and to examine the extent to which activity within the PFC and behavioural indices of inhibition may mediate this relationship.

**Methods:** Sixty-seven adolescents underwent two study sessions five days apart. Sleep and physical activity were measured using wrist-mounted accelerometry; eating habits, substance use and academic achievement were measured by self-report. Prefrontal function was quantified by performance on the Multi-Source Interference Task (MSIT), and task-related brain activity via functional near-infrared spectroscopy (fNIRS).

**Results:** Higher levels of accelerometer-assessed physical activity predicted higher MSIT accuracy scores (*ϐ*= .321, *ρ*= 0.019) as well as greater task-related increases in activation within the right dlPFC (*ϐ*=.008, *SE*= .004, *ρ* =.0322). Frequency of fast-food consumption and substance use were both negatively associated with MSIT accuracy scores (*ϐ*= −.307, *ρ*= .023) and Math grades (*β*= −3.702, *SE*= 1.563, *ρ*= .022) respectively. However, these effects were not mediated by indicators of PFC function.

**Conclusion:** Physical activity and eating behaviors predicted better interference task performance in adolescents, with the former mediated by greater task-related increases in right dlPFC activation. Substance use predicted worse Math grades, however, no other reliable effects of health behaviors on academic outcomes were evident.

## 1.0 Introduction

Strong academic performance depends partially on focused attention, working memory, and planfulness, all of which are partially supported by the lateral and medial subregions of the prefrontal cortex (PFC). Longitudinal (1,2) as well as cross-sectional (3,4) studies have found reliable but modest associations between behavioural measures of these functions and academic performance (5). Health behaviors—including sleep, eating, substance use and physical activity—may all impact the brain in a manner that could affect academic performance among adolescents.

A recent meta-analysis found that sleep restriction has a significant and moderate negative effect on cognitive performance across the lifespan (6). These findings are also supported by imaging studies examining indicators of functional connectivity (7,8). Likewise a high calorie, low nutrient diet has been shown to negatively impact performance on executive function tasks (9,10). Among adolescent girls, a higher Body Mass Index (BMI) correlates with greater impulsivity in response to inhibition tasks as well as reduced activation in the superior frontal gyrus, middle frontal gyrus, ventrolateral prefrontal cortex (PFC), medial PFC (mPFC), and orbitofrontal cortex (OFC; 11). Likewise, there is evidence that substance use and substance use disorders can also be detrimental to brain regions implicated in executive functioning among adolescents. Executive functioning has been shown to be weaker among habitual users of cocaine, amphetamines, cannabis, tobacco and alcohol (12). In addition, youth with a history of alcohol and cannabis use demonstrate less activation in the inferior frontal cortex, but enhanced mPFC response when completing a working memory task (13). The deleterious effects of substance use can potentially persist into early adulthood. When followed for a period of ten years, young adults with a history of alcohol use disorder or a substance use disorder demonstrated poorer performance on verbal and visual learning and memory tasks as well as reduced executive functioning compared to non-users (14). As such, sleep restriction, a poor diet and substance use have all been shown to detrimentally impact academic performance in adolescents (13,15,16). Because each of these lifestyle behaviours have been shown to have a negative relationship with indicators of executive function (e.g., task performance and cortical network engagement), it is plausible to believe that the relationship between each factor and academic achievement may be mediated (in part) through the brain.

In contrast to the apparent adverse effects of the above mentioned health behaviours, both acute and regular physical activity have been shown to enhance some parameters of PFC function and improve task performance on cognitive tasks that tap executive control (17–19). Physical activity training programs have improved the functional and cognitive capacity of older adults (20–22), and these effects appear especially important for brain regions supporting executive control and memory (17,21,23–25). Brain health benefits of physical activity may be present throughout the lifespan, and yet, especially important for adolescents whom may rely on such functions in the academic sphere (17,18,26–28). While the precise pathway through which physical activity influences brain health remains unclear, it is generally thought to increase the production of growth factors critical for synaptic plasticity, angiogenesis and the development of new neuronal architecture, and changes in cerebrovascular dynamics (17). In adolescents, systematic reviews and meta-analyses on the effects of physical activity on executive functions have found net positive effects (29–33) with acute aerobic exercise and in the moderate to vigorous range producing the strongest benefits (31–33).

There is also evidence to support a relationship between greater levels of physical activity and adaptive brain activation during cognitive task performance. A recent study investigating the effects of acute physical activity on the cognitive function of older adults found significantly greater activation in the right and left dorsolateral PFC (dlPFC) post-exercise session during an interference task (22). Although there are very few studies that investigate this topic in children or adolescents, it appears that there is a reliable difference between higher- and lower-fit children. Higher-fit children have been shown to exhibit superior performance on executive performance tasks as well as increased activity in the fronto-parietal regions of the brain (23,34,35). Given the rapid neural development in adolescence, the perceived cognitive benefits of physical activity, both in terms of performance on executive function tasks and through enhanced brain activation, during this critical period may be especially important for the progression of a healthy neurocognitive structure and function into adulthood(36).

Furthermore, the cognitive benefits of exercise could positively impact academic achievement. This “brain benefit” hypothesis postulates that the cognitive enhancements within the PFC could translate into improved academic performance because achievement in school in part relies on executive functions. Currently, the wealth of evidence in support of the relationship between physical activity and academic achievement suggests a null to weak association between the two variables. Systematic and meta-analytic reviews of the literature have shown variable results ranging from null to small positive effects of physical activity interventions (both acute and long-term) on academic performance (29,33,37–41). However, among the studies reviewed there is a large degree of heterogeneity in intervention components assessed, a high degree of variability in the quality of the study designs, and a limited number of studies with sufficient power (29,38,40). More problematic is the inability to achieve blinding (single or double) when assessing physical activity interventions. This also applies to studies involving exercise effects on the brain, which can lead to expectancy effects and therefore an over-estimation of a brain health benefit (in both cognitive testing and functional imaging). In addition, very few randomized trials exist that examine the brain health benefits of exercise in children, and the few that are present have mixed results (23,42). It is possible that overestimations cloud the true effect of physical activity on the brain, and that the cognitive enhancements of physical activity are not potent enough to influence academic achievement in adolescents. Therefore, further investigations into the mediating role of the brain are warranted.

In contrast to the “brain benefit” hypothesis, the removal of physical activity from school curriculums decades ago across North America was often rationalized based on an assumption that physical activity programming competes for time with academic subjects. This perspective posited a negative effect of physical activity on academic performance based on time competition between the two. The “brain-benefit” and “time-competition” perspectives pertaining to physical activity and academic performance suggest a significant relationship, but in opposite directions. It could be that the variation in results from the current body of literature on the topic stems from competition between the two hypotheses. However, if both hypotheses exist, the stronger of the two will determine the net effects (net benefit or net cost). Further research must be done in order to distinguish between the brain benefit and time competition hypotheses.

The emergence of new portable brain imaging technologies—particularly functional Near-Infrared Spectroscopy (fNIRS)—may prove to be more logistically feasible for many types of research studies involving the brain and development and when field settings are required for data collection(43,44). This technology is comparable to functional Magnetic Resonance Imaging (fMRI) in that it measures blood oxygenation parameters for inferring neuronal activity within brain regions (45). Although it has inferior spatial resolution to fMRI, it has better spatial resolution than Electroencephalograms (EEG; 43,44). fNIRS also is less subject to motion artifacts than both EEG and fMRI (46,47) and its portability allows for it to be deployed in field settings more flexibly than any other brain imaging option currently available such as individuals walking outdoors (48), or pilots flying an aircraft (49). Therefore, this technology provides an opportunity to investigate brain activity in a sample of adolescents in a field setting.

The current study examines the relationship between adolescent health behaviours and academic performance as mediated by the brain. Physical activity and sleep hours were assessed using accelerometers, while the other health behaviours (i.e. substance use, fast-food consumption) and academic performance were measured via self-report. The brain health parameters consist of fNIRS measurements of task-related functional activation patterns within subregions of the PFC as well as cognitive interference task performance. It is hypothesized that higher levels of physical activity and greater sleep hours, in addition to less frequent substance use and fast-food consumption will all be associated with greater academic performance, and that this association will be mediated by the brain health parameters.

## 2.0 Material and Methods

### 2.1 Participants and Setting

A sample of 67 adolescent high school students between the ages of 13-18, were recruited for this study. Participants were recruited from one public and three private high schools located in small to medium-sized urban areas. Principals, teachers and/or key members of administration disseminated information and consent materials to the student body. Those students who wished to participate returned their signed consent forms to the administration helping to facilitate the study, or to the student researcher upon the first study session. Demographic features of the sample are presented in Table 1.

**Table 1.** Mean and SDs for the study characteristics.

| Variable | N | % |
| --- | --- | --- |
| Gender |  |  |
| Female | 26.000 | 38.800 |
| Male | 40.000 | 59.700 |
| Age |  |  |
| 13 | 2.000 | 3.000 |
| 14 | 13.000 | 19.400 |
| 15 | 6.000 | 9.000 |
| 16 | 15.000 | 22.400 |
| 17 | 26.000 | 38.800 |
| 18 | 5.000 | 7.500 |
| Substance use (times in past month) |  |  |
| 0 times | 46.000 | 68.700 |
| 1-2 times | 10.000 | 14.900 |
| 3-5 times | 1.000 | 1.500 |
| 6+ times | 10.000 | 14.900 |
| Variable | Mean | SD |
| Fast-food consumption (times in past week) | 1.909 | 1.444 |
| Grades (%) |  |  |
| English | 80.910 | 9.569 |
| Math | 79.300 | 12.076 |
| Average sleep hours | 7.162 | 1.046 |
| Average steps counts | 8914.279 | 3451.436 |
| Average active minutes | 32.391 | 28.456 |
| MSIT % correct responses | 0.883 | 0.056 |

### 2.2 Procedure

The current study was a 5-day prospective observational study with two on-site data collection visits. During each visit, an open period was held during school hours where students could drop in and participate on their own time. This was necessary in order to ensure that data collection took place during free periods and did not impact class time, and that students could participate on-site with minimal inconvenience.

During the first session, students underwent an assent procedure upon presentation of their signed parental consent form. Next a questionnaire consisting of 6 questions pertaining to demographic information (age and gender), eating behaviours, substance use and academic performance was completed. To measure academic performance, participants were asked to report their past years (2018-2019) English and Math grades in percentages. Finally, students were fitted with the fNIRS headband while a cognitive interference task (Multi-Source Interference Task; MSIT; 45) was performed. In total, this first session took approximately 20 minutes per student. Upon completion of the cognitive task, students were given a Fitbit Inspire watch, oriented to its correct usage, and instructed to wear it consistently until the second data-collection period 5 days later.

The second period took place on the following Friday. During this session, students were asked to return their Fitbit and indicate any instances (day, time and duration) when the watch was removed during the period since the first session.

### 2.3 Demographics, health behaviours, and academic performance

Participants were asked to report their age and gender. In addition, students were asked “how many times have you eaten “fast-food” (e.g. McDonalds, Burger King, etc.) in past week?” as a measure of calorically dense food consumption. Participants were also be asked “how many times have you experimented in the past month with substances (e.g., alcohol, cannabis, other)?”and responded using a scale ranging from 0,1-2, 3-5, 6+. To measure academic performance, students were asked “What was the final grade that you received last year (2018-2019) in Math class?” and “What was the final grade that you received last year (2018-2019) in English class?” Students were then able to indicate their English and Math grade in percentage.

### 2.4 Multi-Source Interference Task

Participants completed the MSIT as a measure of response inhibition (66,67). For this task, each trial consisted of three numbers that were horizontally aligned in the centre of a black computer screen in bold white 50pt font. A “+” sign was presented between trials and in the centre of the screen during a 1.75s inter-trial interval. The numbers corresponded to the “1, 2 and 3” numbered computer keys, and participants were instructed to indicate the unique number in the triplet by pressing the corresponding key using their dominant hand.

Control and interference trials differed by the type of distractor used as well as the position of the target numbers in relation to their location on the keyboard. In control trials, the target number always matched their location on the keyboard, and the distractors were never used as targets. During interference trials, the target number never matched its position on the keyboard and the distractors were also targets. During the initial orientation to the MSIT task, participants were asked to respond as quickly and accurately as possible in response to each number stimulus. A 1.5 -minute practice trial of 48 trials initiated the task (50,51). Participants then completed 4 blocks of 24 control and interference trials for a total of 96 trials of each type. There was a fixed order of trials within each block, but blocks were counterbalanced between participants (i.e. either CICICICI or ICICICIC). A 30 second rest period with a fixation cross was included at the beginning of the first block and at the end of the last block (50,51).

### 2.5 Functional Near-Infrared Spectroscopy

fNIRS is an optical neuroimaging technique which non-invasively measures activation of the cortex using near-infrared (NIR) light (46,47). In order to measure regional activation, fNIRS relies on metabolic related increases local arteriolar vasodilation, which in turn produces an increases in oxygenated hemoglobin (OxyHb) as well as a (slightly time lagged) relative decrease in deoxygenated hemoglobin (DeoxyHb; 43). Because hemoglobin is the main chromophore that absorbs NIR light and does so differently when oxygenated (>800nm) versus deoxygenated (<800nm), fNIRS can utilize the spectroscopic features of hemoglobin in order to infer regional brain activation (46).

For this study, 16-prefrontal cortical regions were monitored using fNIR Devices 203C imaging system during the MSIT task. The system collected data at 10Hz sampling rate for both 730nm and 850nm as well as ambient light intensity and two additional short distance channels. The dependent variable was the change in OxyHb between MSIT control and interference trials in each target area from 2 seconds to 8 seconds and relative to baseline. A rest period for 20 seconds preceded measurements. Four regions of interests (ROI) were identified, corresponding to conceptually important subregions of the prefrontal cortex. Channels 3,4 and 6 make up the left dlPFC (L-dlPFC); 13,14 and 15 the right dlPFC (R-dlPFC); 7,8 the left mPFC (L-mPFC) and 9,10 the right mPFC (R-mPFC). Raw light intensities in the 730 nm and 850 nm wavelengths as well as ambient light intensities were recorded using the COBI Studio software (52). Each participant’s data was checked for any potential saturation (when light intensity at the detector was higher than the analog-to-digital converter limit) and motion artifact contamination by means of a coefficient of variation based assessment, sliding window motion artifact rejection (53). A low-pass finite impulse response filter with an order of 100 and cut-off frequency of 0.1Hz was designed using Hamming window and was subsequently applied to the light intensity measures to attenuate high frequency noise and physiological nuisance signals (i.e. heartbeat, respiration). Using the modified Beer-Lambert law, raw light intensities were then converted into OxyHb and DeoxyHb concentrations (54). fNIRS data for eight training blocks (4 control and 4 interference) of 42 seconds were extracted using based on synchronization markers. The hemodynamic response at each optode was baseline corrected using the linear subtraction of the signal level at the beginning of each trial block, and finally averaged across time for each trial block to provide a mean hemodynamic response at each channel for each block.

### 2.6 Accelerometry

At the conclusion of the first 20-minute session, each participant was provided a Fitbit Inspire watch to wear until the second session on the following Friday. Each Fitbit watch was embedded with an triaxial accelerometery sensor that measured linear acceleration along three orthogonal axes (X, Y and Z) and can detect movement including gravity (55). The accelerometry sensor was used to determine active minutes and sleep hours. Students were instructed to wear the Fitbit consistently Monday to Friday, day and night. Each Fitbit Inspire watch was linked to a Fitbit account so that physical activity and sleep data could be accessed remotely and recorded. The Fitbits were synced using the Fitbit app once during the first and last sessions. During the final session, students were asked to drop in and return their Fitbit watch and specify whether or not the watch was removed during the week using a weekly calendar.

### 2.7 Statistical analysis

All statistical analyses were conducted using SPSS. The Explore subcommand in SPSS was used to generate Boxplots and distributional statistics; together these were employed to assess skewness and kurtosis for each individual variable, and to any extreme outliers that may be present. One extreme outlier was removed from the fast-food consumption variable and three extreme outliers were removed from the % correct MSIT responses variable. English grades, the % correct MSIT responses, the mean MSIT RT and MSIT SD, were subjected to winsorization.

To calculate the fNIRS indicators of oxygenation in each ROI the mean change in OxyHb was calculated for the interference and control blocks of the MSIT separately and for each channel. These mean values of oxygenation were then transformed into Z scores and the mean of the Z scores were calculated for all channels making up each ROI. All ROI aggregates were then subjected to winsorization.

Hierarchical linear regression models were employed in order to examine the relationship between each lifestyle behavior (i.e., physical activity, sleep hours, fast-food consumption frequency and substance use frequency) and MSIT performance (% correct responses) while controlling for age. The PROCESS macro was utilized run moderated regression analyses in order to examine the moderating effect of gender and BMI on the above models (i.e., all lifestyle factors and MSIT % correct responses). Multiple mediation models were then utilized to assess the potential mediating effect of the brain health parameters (MSIT indicators, fNIRS ROI oxygenation) while controlling for the % correct responses. Final conditional process models assessed whether the above multiple mediation models differed based on gender, age or BMI. An estimate of BMI was calculated by the Fitbit watches upon the first day of wear, and relied on physical characteristics (year of birth, height, weight, sex) supplied by the participants.

## 3.0 Results

Initial predictive models were fitted using multi-level modelling of within-person effects pertaining to trials within blocks and blocks within task. Modelling these nested effects did not significantly improve model fit, and so the primary analyses presented below utilized data averaged across trials and blocks of the same type. This enabled the use of multiple mediation models, using an ordinary least squares regression approach, which forms the primary analytic approach in the sections below. Given the focus of the study on executive control and evaluative processes, our functional imaging analyses were primarily focussed on oxygenated hemoglobin levels in interference blocks. During signal processing, each individual task trial was divided into a 2 second baseline and 8 second sampling epoch, and so OxyHb levels described below represent average changes (increases normally) in OxyHb from each of these local baselines during each individual task trial.

Initial analyses show lifestyle behaviors as predictors of interference task performance (% correct MSIT responses). These are followed by multiple mediation models testing simultaneous mediational effects of a given target behavior (e.g., activity, sleep, etc.) on an academic performance outcome (e.g., Math grades or English grades) through all brain primary health parameters (e.g., MSIT reaction mean reaction times; MSIT reaction time variability; fNIRS parameters), while controlling for MSIT task performance. Subsequent conditional processed models explored the extent to which the multiple mediation models were moderated by age, gender and BMI. fNIRS channels were combined into neuroanatomically relevant ROIs, corresponding to the L-dlPFC, R-dlPFC, L-mPFC, and R-mPFC. All multiple mediation and conditional process models focussed on these four regions of interest.

### 3.1 Lifestyle predictors of interference task performance

Means and standard deviations (SD), as well as N and % for categorical variables, for all sample characteristics and primary study variables are presented in Table 1. The majority of students were aged 16-17 (61.2%) and Male (59.7%).

To examine the extent to which MSIT performance was predicted by each target lifestyle behavior, behavior-specific regressions were run using age as a covariate and each lifestyle behavior as a predictor. Findings are presented in Figure 1. Both fast-food consumption and accelerometer-assessed active minutes were significant predictors of MSIT performance. Specifically, more average daily minutes of accelerometer-assessed physical activity predicted greater % correct MSIT responses (*β*= .321, *ρ*= .019); likewise, less frequent fast-food consumption in the previous week predicted significantly greater % correct MSIT responses (β= −.307, *ρ*= .023). A reduced model predicting MSIT performance was created containing only average active minutes and fast-food consumption. This reduced model accounted for 24% of the variability in MSIT performance (∆*R^2^* = .244, *ρ* < .001). In the reduced model, higher average daily minutes of accelerometer-assessed physical activity predicted greater % correct MSIT responses (β=.382, *ρ*= .004). Likewise, less frequent fast-food consumption in the past week predicted significantly higher % correct MSIT responses (β= .429, *ρ*= .001).

**Figure 1.**
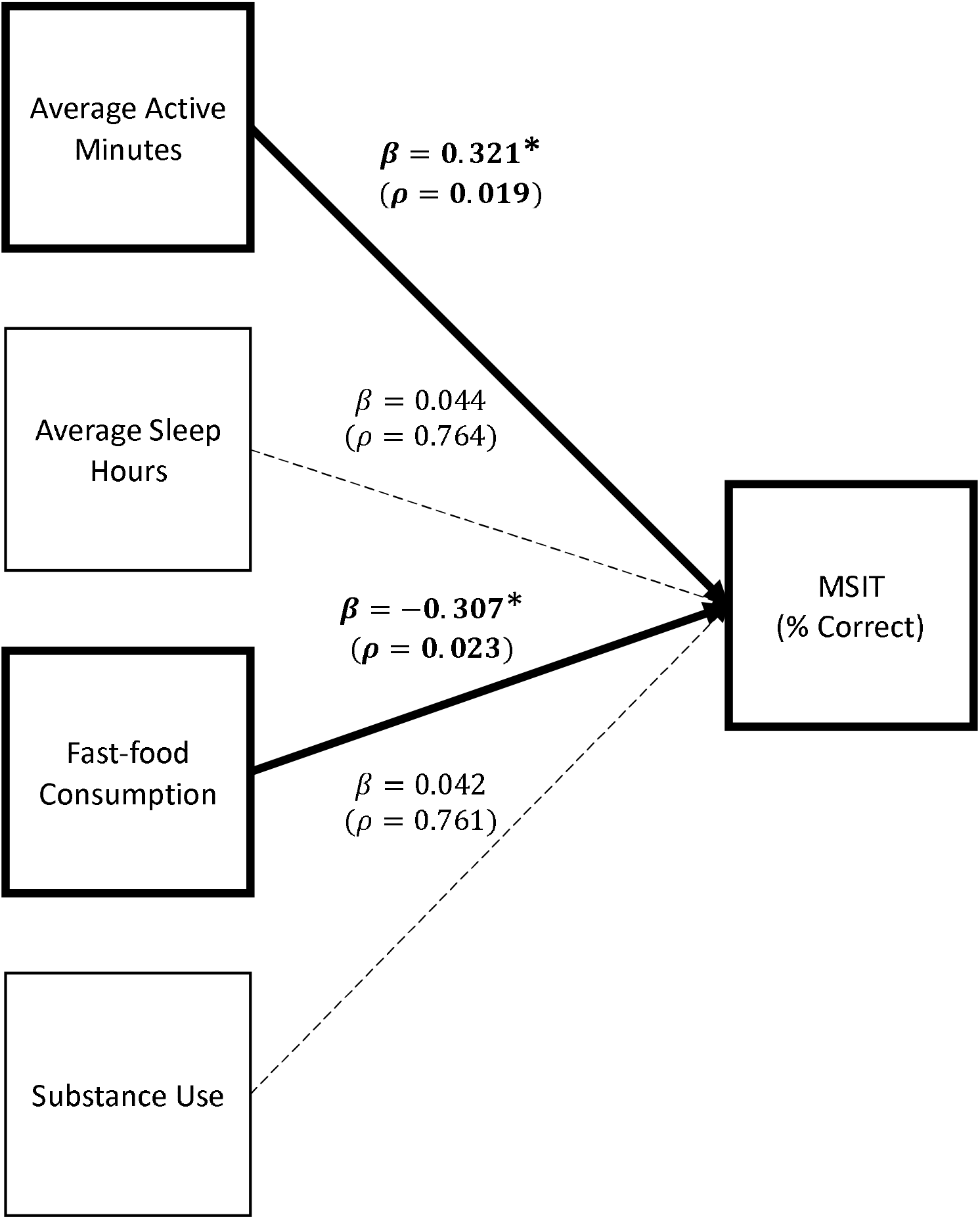
Lifestyle behaviors predicting MSIT performance, controlling for age.

To determine whether the strength of association between each predictor and MIST performance differed for male and female participants, moderated regression analyses were performed separately for each target behavior. Gender was a significant moderator of the relationship between active minutes and MSIT performance (∆*R^2^* = .077, *F*= 4.939 (1, 54), *ρ* =.031), such that active minutes had a significant effect on MSIT performance for females (*β* = 1.018, *SE*= .412, *ρ*= .017) but not for males (*β* =.041, *SE*= .155, *ρ*=.793). The corresponding effect sizes were .061 for males and .384 for females (Figure 2). Gender was not a significant moderator of the relationship between fast-food consumption and MSIT performance (∆*R^2^* = .011, *F*=.699 (1, 59), *ρ* =.407). There was also no significant moderating effect of gender on the relationship between sleep (∆*R^2^* = .043, *F*= 2.194 (1, 47), *ρ* =.145) or substance use (∆*R^2^* = .000, *F*= .010 (1, 59), *ρ* = .919) and MSIT performance.

**Figure 2.**
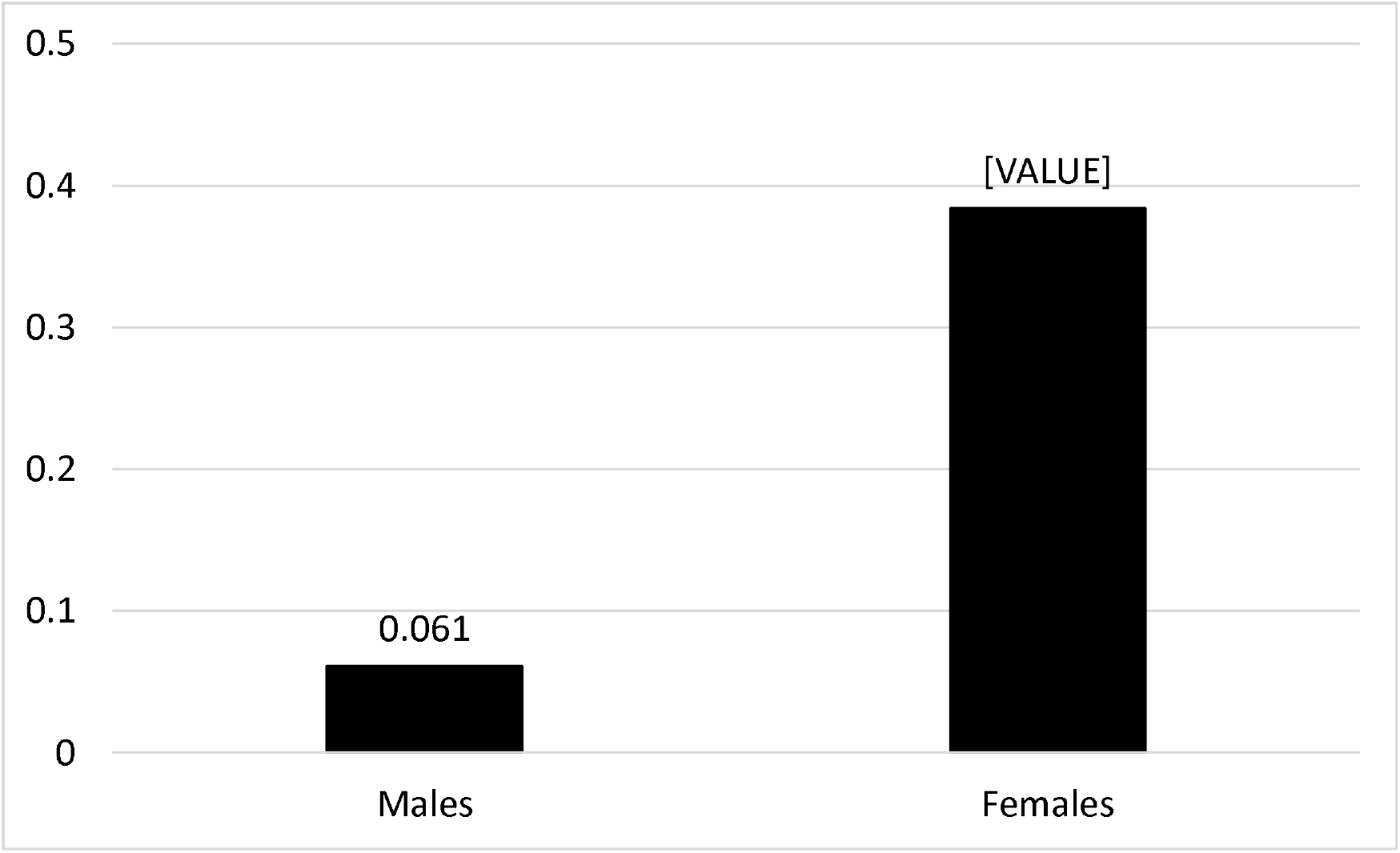
Effect sizes for average active minutes predicting MSIT performance (% correct)

Additional moderated regression analyses were performed to determine whether the strength of association between each predictor and MIST performance differed by body composition (quantified by BMI). BMI was calculated using percentile-based age and gender-specific cut-offs recommended by Centers for Disease Control and Prevention for children and adolescents (56). Results indicated that BMI was indeed a significant moderator of the relationship between substance use and MSIT performance (∆*R^2^* = .127, *F*= 8.916 (1, 59), *ρ* =.004). The corresponding effect sizes were .119 for those for those whose BMI fell within the obese range and −.095 for non-obese (Figure 3). BMI was not a significant moderator of the relationship between active minutes (∆*R^2^* = .008, *F*= .492 (1, 55), *ρ* =.486), fast-food consumption (∆*R^2^* = .019, *F*= 1.171 (1, 58), *ρ* =.284), or average sleep hours (∆*R^2^* = .027, *F*= 1.384 (1, 48), *ρ* =.245) and MSIT performance.

**Figure 3.**
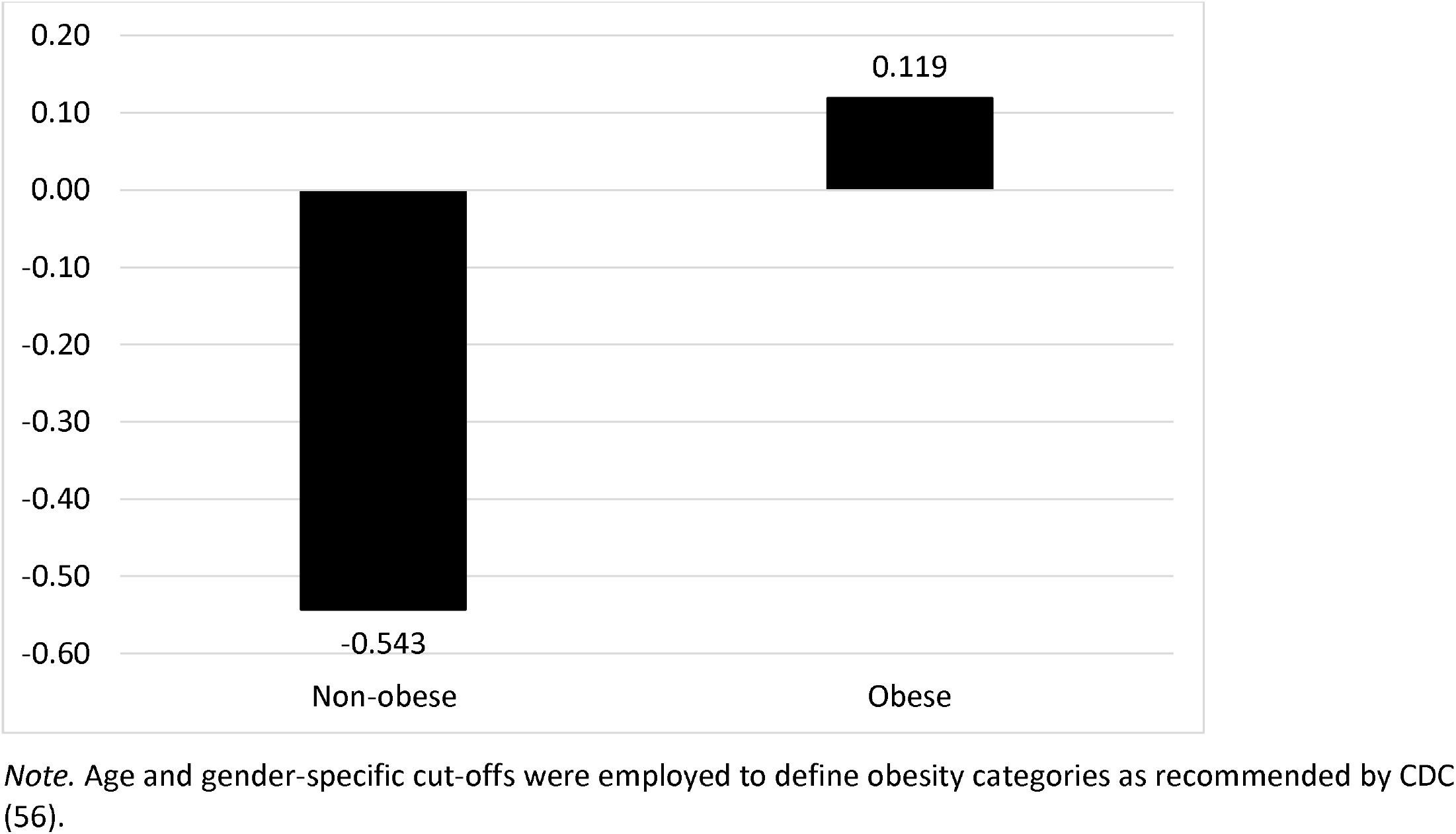
Effect sizes for substance use predicting MSIT performance (% correct)

### 3.2 Multiple mediation models

In order to examine mediational processes predicting academic achievement from target lifestyle behaviors via candidate brain health mediators (MSIT parameters, fNIRS ROI), multiple mediation models were fitted using the PROCESS Macro in SPSS. This analysis was completed separately for each lifestyle behaviour (i.e., average sleep hours, average active minutes, fast-food consumption and substance use) and each academic outcome variable (i.e., English and Math grades), while controlling for % correct MSIT responses.

#### 3.2.1 Math grades

##### 3.2.1.1 Physical activity multiple mediation model

Figure 4 depicts the multiple mediation model predicting Math grades from average active minutes through brain health parameters. There was a significant effect of average active minutes on R-dlPFC OxyHb (*β* = .008, *SE*= .004, *ρ* =.032), but no effect of average active minutes on L-dlPFC OxyHb (*β* = .003, *SE*= .003, *ρ* = .295), L-mPFC OxyHb (*β* = .003, *SE*= .003, *ρ* =.401), or R-mPFC OxyHb (*β* = −.003, SE= .003, *ρ*= .266). Figure 5 a) displays a heat map showing channel specific effects of average active minutes, and figure 5 b) shows areas of significant activation overlaid on a 3-dimentional anatomical brain. There was also no direct effect of average active minutes on Math grades (*β*=.106, SE=.070, *ρ*= .139), and no effect of average active minutes on either the MSIT mean RT (*β*= −.006, SE=.005, *ρ*= .184), or on the MSIT SD (*β* = .002, SE = .004, *ρ* = .676). None of the brain health parameters were significant predictors of Math grades.

The indirect effect of average active minutes on Math grades through R-dlPFC OxyHb was not significant; the upper and lower bound for the 95% confidence interval for the indirect effect included zero (est.= .011 (*SE*= .025); *CI_LL_* = −.039, *CI_UL_* = .064), suggesting a null mediational effect. None of the other indirect effects involving brain health parameters were significant (See supplementary materials).

**Figure 4.**
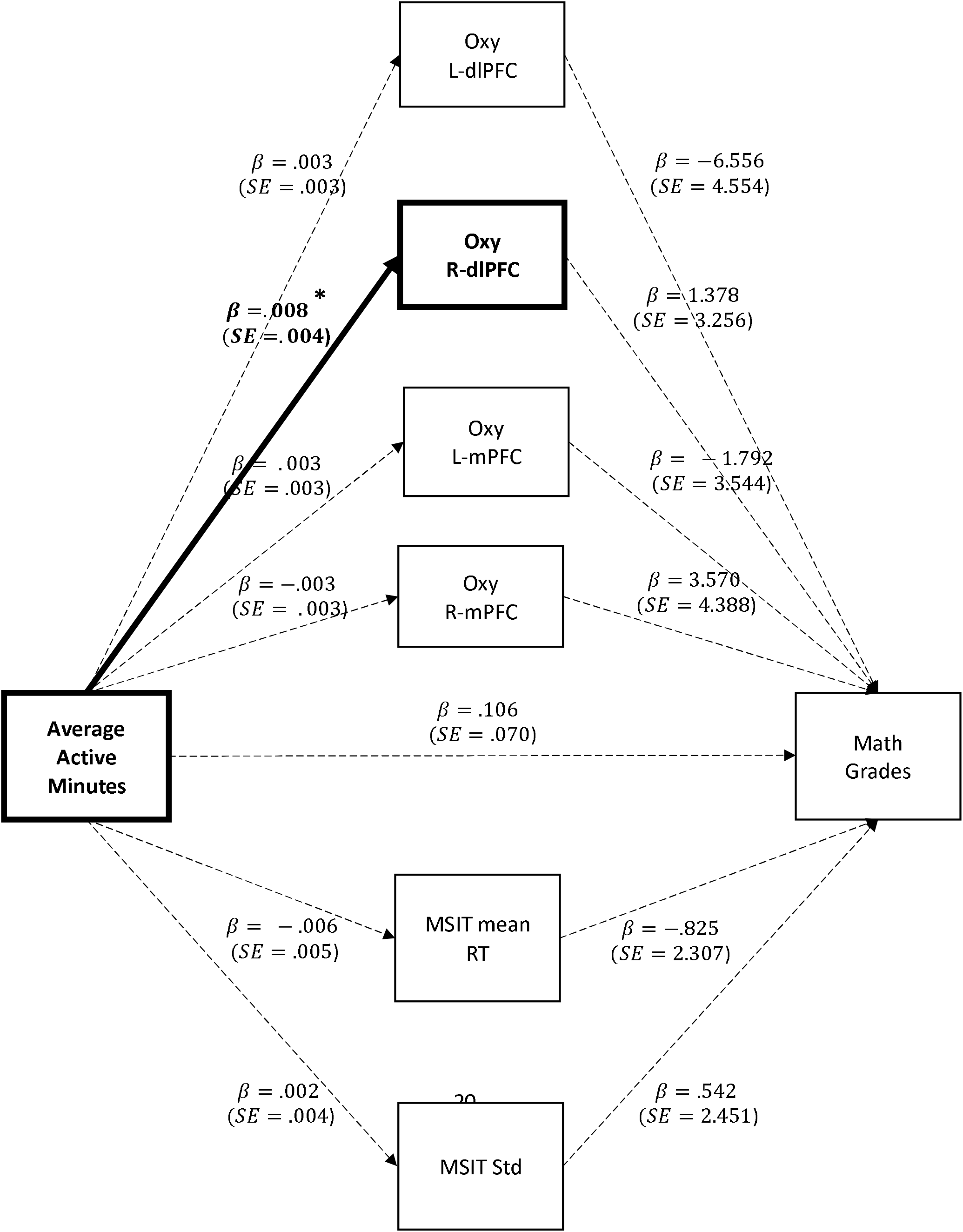
Multiple mediation model predicting Math grades from accelerometer-assessed active minutes of physical activity through brain health parameters, controlling for MSIT % correct responses.

**Figure 5.**
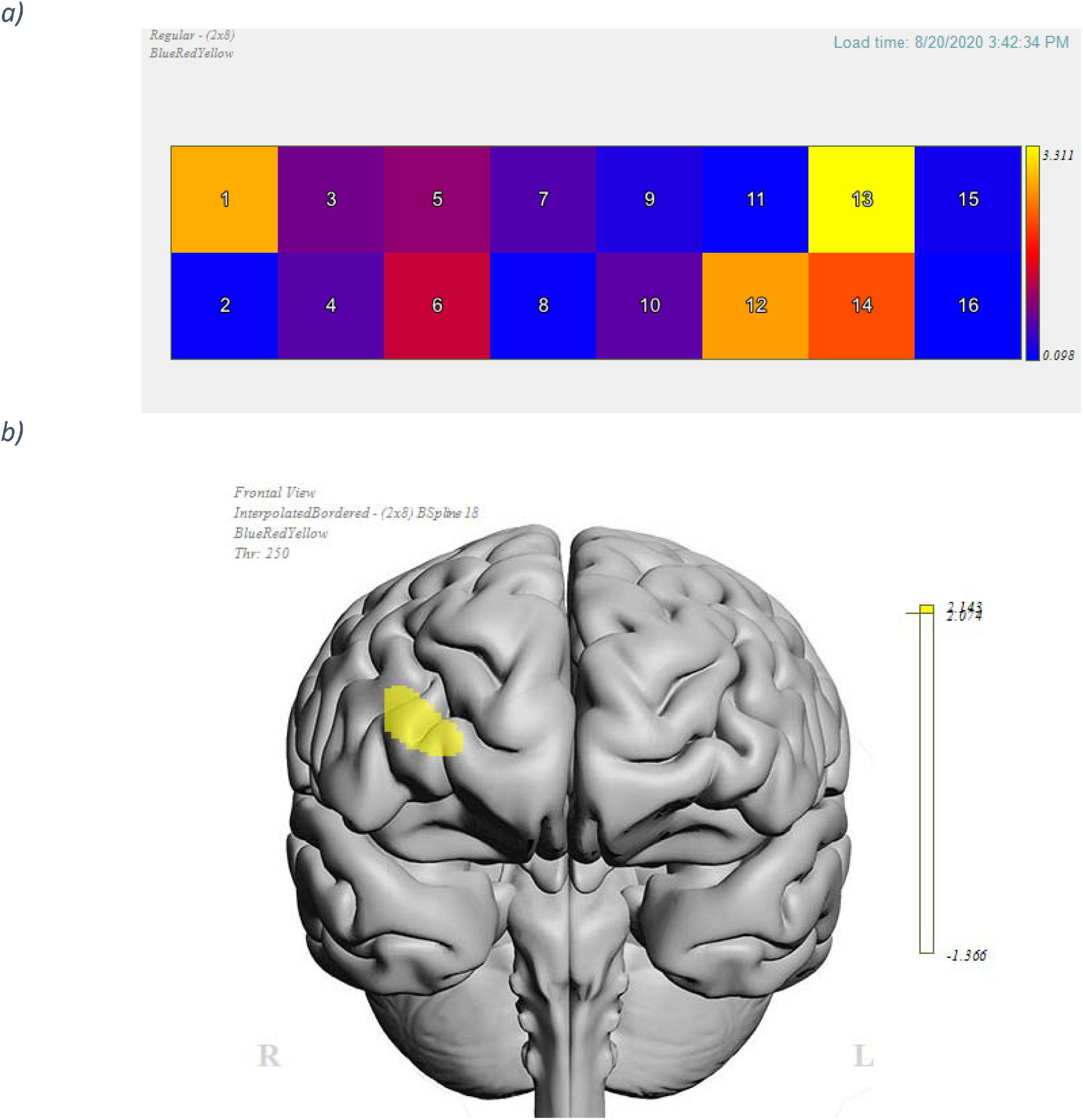
a) Heat map portraying F statistics for the effect of average active minutes on mean OxyHb levels in each channel and b) anatomical position of significant effects.

##### 3.2.1.2 Substance use multiple mediation model

Figure 6 depicts the multiple mediation model predicting Math grades from substance use through brain health parameters. There was a significant direct effect of substance use on Math grades; specifically, more frequent substance use in the past month predicted significantly worse math grades (*β*= −3.702, SE= 1.563, *ρ*= .022). There was no effect of substance use on either the MSIT mean RT (*β*=.017, SE=.112, *ρ*= .883), or on the MSIT SD (*β*=.159, SE=.099, *ρ* = .113). There was also no significant effect of substance use on LdlPFC OxyHb (*β*= −.015, SE=.064, *ρ* = .813), R-dlPFC OxyHb (*β*=−.107, SE=.090, *ρ* = .240), L-mPFC OxyHb (*β*=−.106, SE=.074, *ρ* = .176), or R-mPFC OxyHb (*β*=−.036, SE=.067, *ρ*= .630). None of the brain health parameters were significant predictors of Math grades, and none of the indirect effects involving brain health parameters were significant (See supplementary materials).

**Figure 6.**
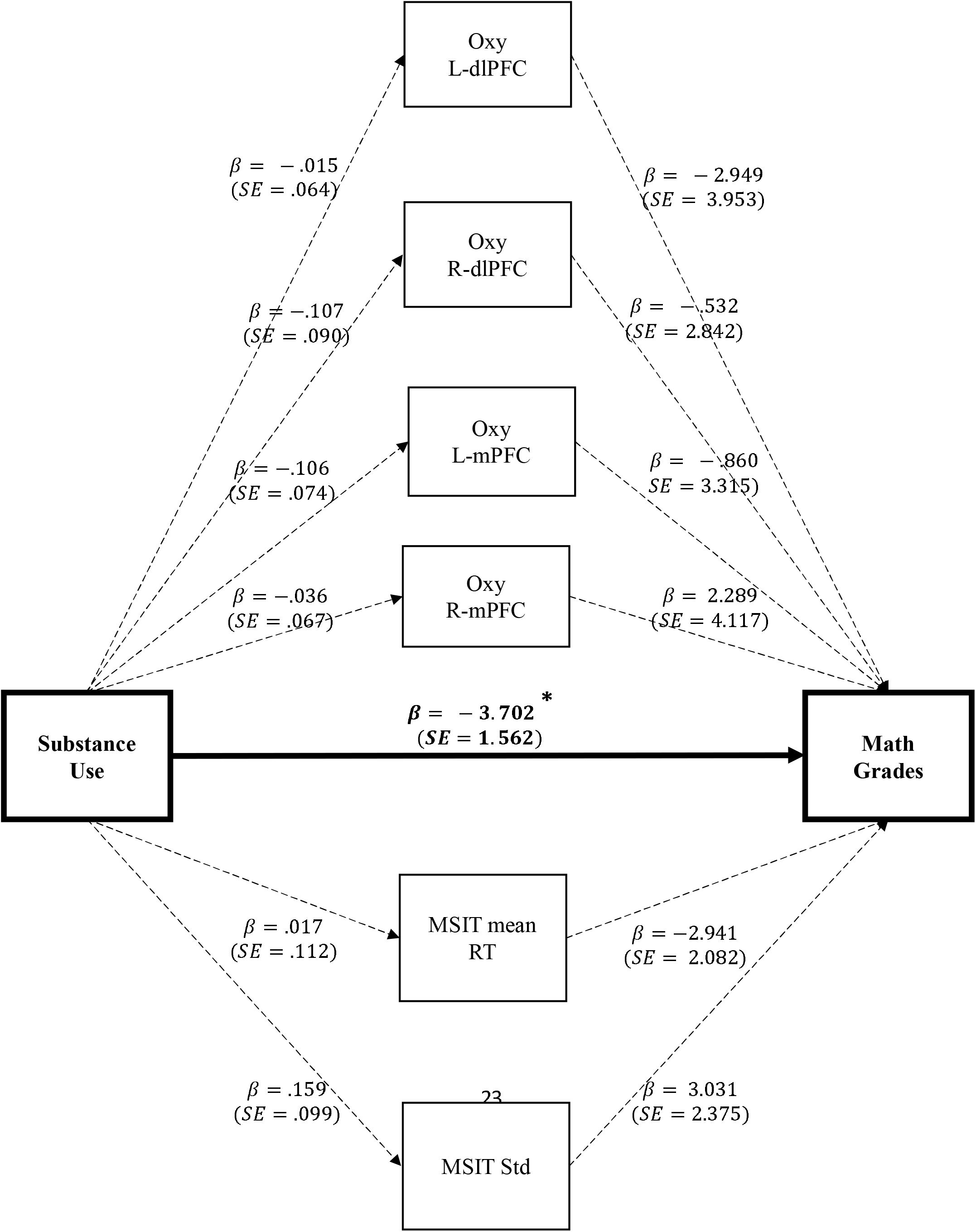
Multiple mediation model predicting Math grades from self-reported substance use through brain health parameters, controlling for MSIT % correct responses.

##### 3.2.1.3 Fast-food consumption multiple mediation model

A multiple mediation model predicting Math grades from fast-food consumption through brain health parameters found was no direct effect of fast-food consumption on Math grades (*β*= −.882, SE=1.297, *ρ*= .500), and no effect of fast-food consumption on either the MSIT mean RT (*β*= −.037, SE= .088, *ρ*= .679), or on the MSIT SD (*β*= .128, SE=.078, *ρ* = .109). There was also no significant effect of fast-food consumption on LdlPFC OxyHb (*β*=.008, SE=.050, *ρ* = .874), R-dlPFC OxyHb (*β*= −.061, SE= .072, *ρ* = .406), L-mPFC OxyHb (*β*= .021, SE=.060, *ρ* = .731), or R-mPFC OxyHb (*β*=.008, SE=.053, *ρ*= .885). None of the brain health parameters were significant predictors of Math grades, and none of the indirect effects involving brain health parameters were significant (See supplementary materials).

##### 3.2.1.4 Average sleep hours multiple mediation model

A multiple mediation model predicting Math grades from average sleep hours through brain health parameters found no direct effect of average sleep hours on Math grades (*β*=−.742, SE=1.807, *ρ*= .684), and no effect of average sleep hours on either the MSIT mean RT (*β*=.090, SE=.129, *ρ*= .488), or on the MSIT SD (*β*=.025, SE=.110, *ρ* = .823). There was also no significant effect of average sleep hours on L-dlPFC OxyHb (*β*=.065, SE=.074, *ρ* = .385), R-dlPFC OxyHb (*β*=.167, SE=.104, *ρ* = .114), L-mPFC OxyHb (*β*= −.046, SE=.081, *ρ* = .577), or R-mPFC OxyHb (*β*=−.077, SE=.081, *ρ*= .346). None of the brain health parameters were significant predictors of Math grades, and none of the indirect effects involving brain health parameters were significant (See supplementary materials).

#### 3.2.2 English grades

##### 3.2.2.1 Physical activity multiple mediation model

The above multiple mediation models were repeated with English grades as the outcome variable. Figure 7 depicts the multiple mediation model predicting English grades from average active minutes through the brain health parameters. There was a significant effect of average active minutes on R-dlPFC OxyHb (*β* = .008, *SE*= .004, *ρ* =.032), but no effect of average active minutes on L-dlPFC OxyHb (*β* = .003, *SE*= .003, *ρ* = .295), L-mPFC OxyHb (*β* = .003, *SE*= .003, *ρ* =.401), or R-mPFC OxyHb (*β* = −.003, SE= .003, *ρ*= .266). There was also no direct effect of average active minutes on English grades (*β*=−.070, SE= .055, *ρ*= .659), and no effect of average active minutes on either the MSIT mean RT (*β*= −.006, SE=.005, *ρ*= .184), or on the MSIT SD (*β* = .002, SE = .004, *ρ* = .676). None of the brain health parameters were significant predictors of English grades.

The indirect effect of average active minutes on English grades through R-dlPFC OxyHb was not significant; the upper and lower bound for the 95% confidence interval for the indirect effect included zero (est.= .020 (*SE*= .027); *CI_LL_* = −.023, *CI_UL_* = .081), suggesting a null mediational effect. None of the other indirect effects involving brain health parameters were significant (See supplementary materials).

**Figure 7.**
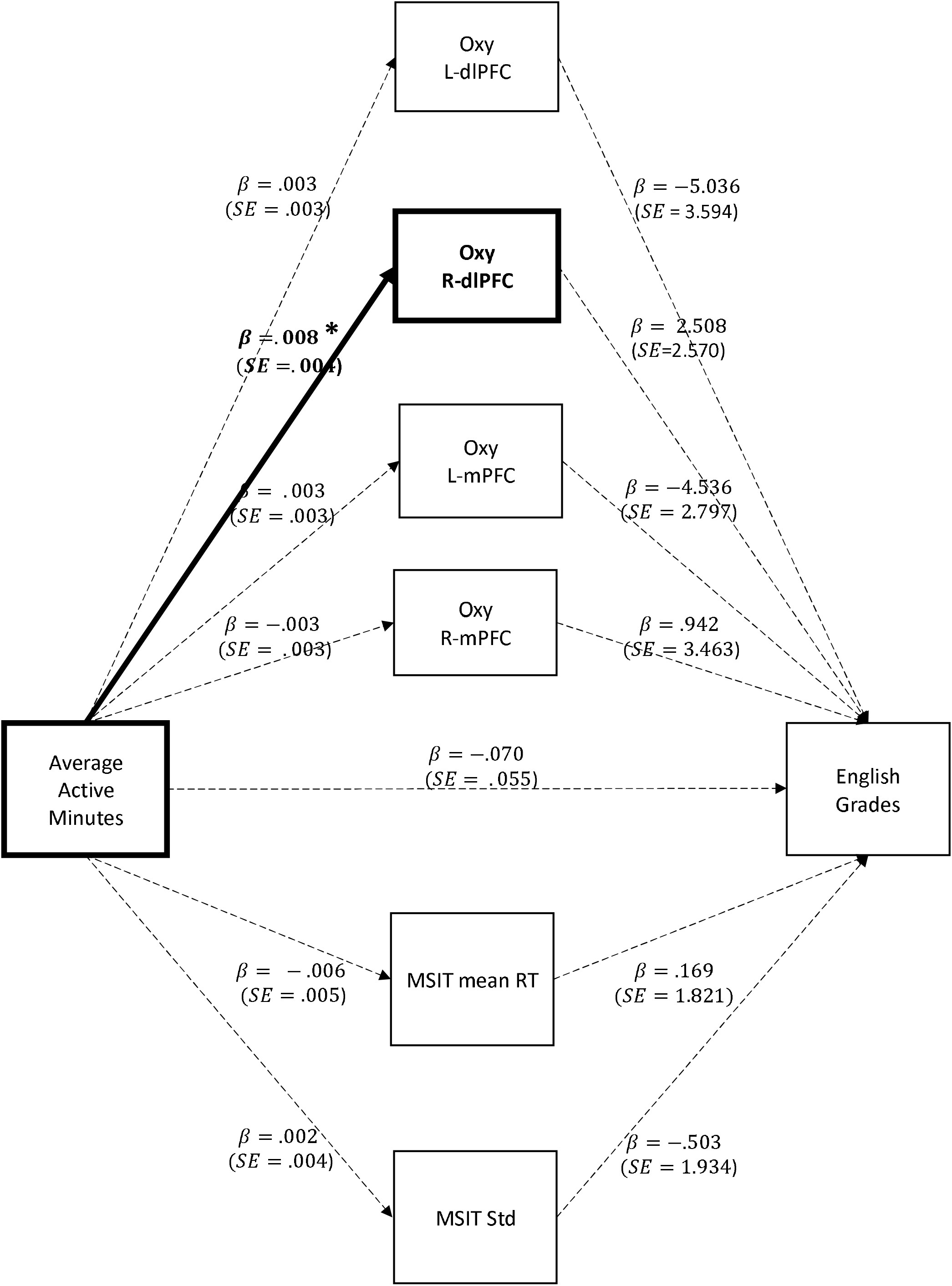
Multiple mediation model predicting English grades from accelerometer-assessed active minutes of physical activity through brain health parameters, controlling for MSIT % correct responses.

#### 3.3 Conditional process models

A set of conditional process models were tested to examine the extent to which lifestyle behaviour associations with academic performance mediated through brain health variables might be conditional upon age, gender and BMI. A significant moderation effect emerged regarding BMI on the indirect effect of average active minutes on English and Math grades through MSIT SD (∆*R^2^* = .086, *F*=6.318 (1, 45), *ρ* =.016). Specifically, average active minutes had a stronger effect on the MSIT SD for those in the highest BMI category (*β*= .431, *SE*= .192, *ρ*= .030). The corresponding effect sizes were .112 for those in the obesity range (≥ 95^th^ percentile) and −.095 for all others (<95^th^ percentile; Figure 8). All additional models produced null moderated mediational effects (See supplemental material).

**Figure 8.**
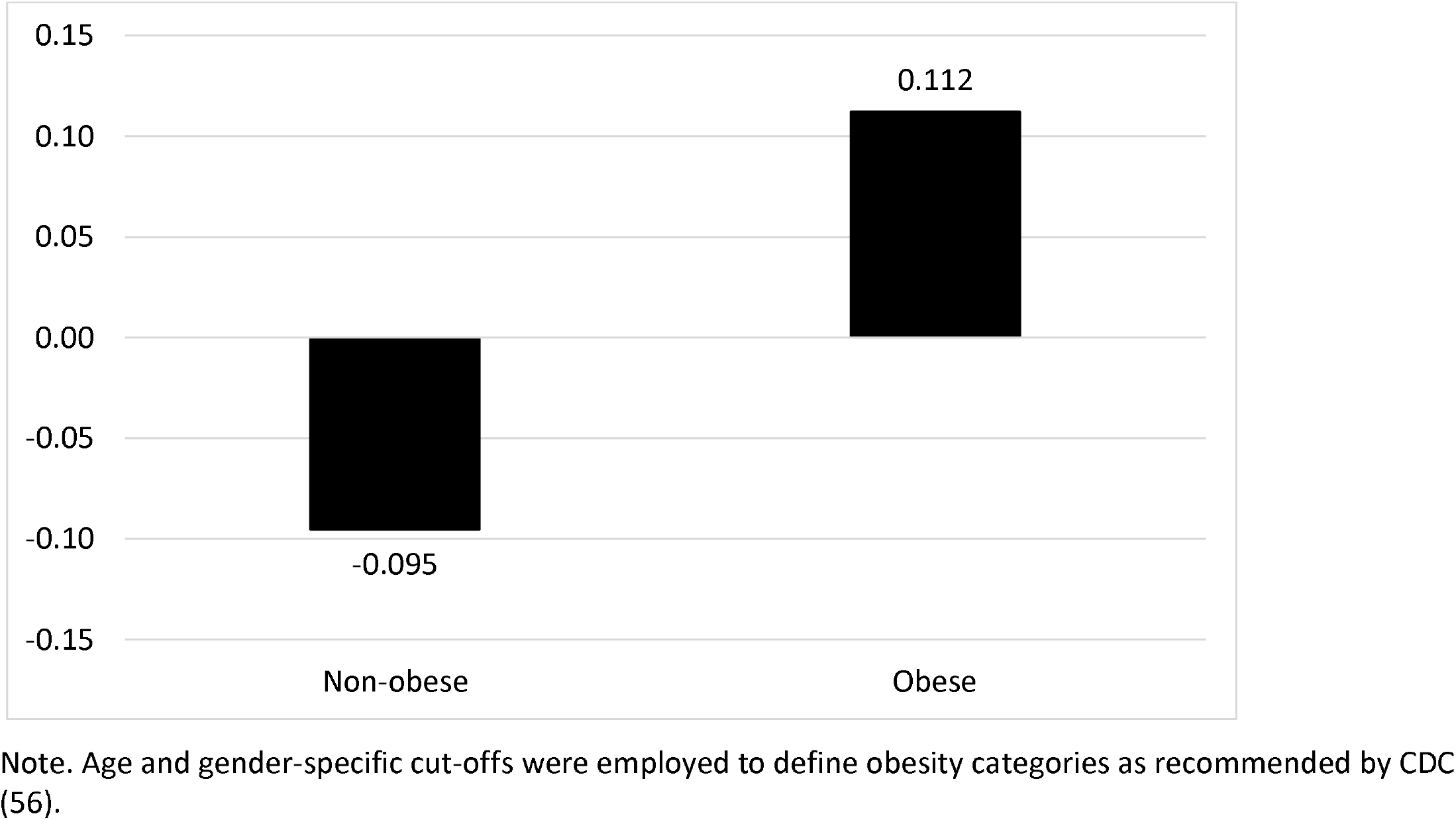
Effect sizes for active minutes predicting MSIT SD.

### Discussion

The purpose of this study was to examine the extent to which the relationships between health behaviors and academic performance might be mediated by brain health parameters in a sample of adolescents. A prospective observational design was employed, utilizing a combination of accelerometery and self-reported measures of health behaviors as well as fNIRS and MSIT quantifications of brain health. Findings revealed that higher levels of accelerometer-assessed physical activity, as well as less frequent fast-food consumption both independently predicted significantly better interference task performance. Higher levels of physical activity were associated with larger increases in task-related oxygenation in the right dlPFC during interference blocks, and relative to baseline. There were, however, no significant links between physical activity or fast-food consumption and academic achievement, either directly or mediated by brain health parameters.

A multiple mediation approach was used to investigate the relationship between each lifestyle predictor and academic outcomes, as mediated through all brain health mediators (task performance and functional imaging parameters). Fast-food consumption was associated with MSIT performance, but eating habits were not significantly associated with any other MSIT indicator (i.e., MSIT mean RT and SD) or oxygenation in any ROI. Higher levels of self-reported substance use were associated with poorer performance in Math. Finally, average sleep hours were not significantly associated with either of the academic outcomes, or indirectly associated with the brain health parameters.

Conditional process (i.e., moderated mediation) models found a significant moderating effect for BMI on the indirect effect of average active minutes on English and Math grades through reaction time variability on the interference task. Initial models also suggested that BMI was a significant moderator in the relationship between substance use and task performance, where performance on the interference task was better for those whose BMI fell within the obese range. However, there was no moderating effect of BMI in the relationship between substance use and task mean reaction time, or reaction time variability when utilizing the conditional process models.

Although there was no direct effect of physical activity on either English or Math achievement, greater levels of physical activity were associated with better performance on the cognitive interference task and higher levels of neuronal activation in the R-dlPFC during the interference task. This is consistent with a wealth of experimental findings, which have found cognitive benefits of physical activity across age ranges (17,21,28), when investigating the effects of activity on task related performance using various interference tasks (e.g., Stroop, Flanker; 58,110,111), and when utilizing various neuroimaging techniques (18,22,23,34). Although stronger activation in the L-dlPFC is commonly associated with interference tasks, the association between active minutes and oxygenation in the R-dlPFC still supports the notion that physical activity does promote greater activation within the PFC.

Contrary to hypotheses, there was no direct association between physical activity and either indicator of academic achievement. It is possible that such activity-induced brain benefits were simply not strong enough to induce changes in academic achievement. Meta-analyses on the topic have shown a small effect of physical activity on academic performance in children (38,58), and so a high statistical power may be required to detect a translatable effect. Alternatively, academic performance is an outcome that is highly multi-determined, such that it relies on the cooperation of many cognitive processes (3,59) and is a construct that can be influenced by many external factors, including socioeconomic status, race and gender (116,117). Consequently, activity may only marginally impact academic performance because the total universe of influences is so expansive. Likewise, the time-competition hypothesis, which implies a negative association between physical activity and academic performance, or an interaction between the brain benefit and time competition hypotheses, could cause this pattern of findings such that any brain health benefits of activity could be offset by the time detracted from academic pursuits. The pattern of data in the current study is most consistent with this latter interpretation.

Interestingly, performance on the cognitive interference task was moderated by gender, such that the effect of physical activity on task performance was stronger for females than for males. Sex has previously been shown to moderate the effects of physical activity effects on the brain, but primarily in older adults (62). It has been hypothesized that differences between females and males sex steroid hormones could impact cognition and that the hormonal environment during pubertal development may have enduring effects on both the structure and function of the brain from the adolescent developmental period and onwards (62,63). This may explain why the current findings suggest that such sex differences may extend to lower age ranges. It could be argued from this perspective that greater emphasis on physical education for females could be undertaken in schools, if any associated time competition effects could be offset.

In addition to the effects of physical activity on task performance, more frequent fast-food consumption was associated with poorer performance on the cognitive interference task. However, the correlational nature of this study does not allow for interpretations of the temporal relationship between these two variables. Previous studies have identified a negative relationship between poor dietary habits and reduced executive functioning (10,64), but experimental studies involving transient up- or down-regulation of the lateral PFC have produced causal changes in food consumption, which supports the alternative temporal relationship (65,66). Notwithstanding, the results of this study do emphasize importance of dietary quality on executive functioning in this age group.

Furthermore, greater substance use was found to predict poorer Math grades. This is consistent with hypotheses, and prior findings that substance use among adolescents negatively impacts academic achievement (16,67). Substance use experimentation and substance use disorders often emerge in adolescents (68) and this variable was the only significant predictor of academic achievement, high schools should be especially aware of the impact of substance use when tailoring preventative initiatives.

Limitations of this study include the observational research design, which limits the extent to which causal effects can be identified. Likewise, some variables were studied with temporal lags but not others, making directionality inferences challenging in the latter. In addition, the small sample size may have resulted in reduced statistical power to detect effects, which could have impacted the ability to detect significant associations. Furthermore, the field setting introduced noise into the fNIRS measurements, and the use of self-reported academic achievement variables may have been subjected to social desirability bias, which in turn may have contributed to and lower quality data.

Key strengths of this study included the use of objective measures of brain activation, physical activity and sleep; the latter two in particular are thought to be more reliable and valid than self-report measures (45,69). In addition, the use of both fNIRS imaging and a standardized cognitive task allowed for the derivation of several indicators of brain health, both in terms of behavioural markers of executive function and task related activation in the PFC. Furthermore, the field setting allowed for direct recruitment of students in a naturalistic environment. Finally, there are relatively few studies that employ a brain imaging protocol in order to investigate how lifestyle behaviours impact academic performance through brain health parameters in adolescents and so this study fills a gap in the current literature on the subject.

### 3.4 Conclusion

Utilizing a sample of adolescents, this study aimed to identify to what extent the relationship between lifestyle behaviours and academic performance was mediated by brain health. Although there was no direct association present between accelerometer-assessed physical activity and either English or Math grades, greater active minutes did have a positive effect on interference task performance and on lateral PFC recruitment. The effect of active minutes on cognitive task performance was also moderated by gender, such that females experienced a greater cognitive benefit of physical activity compared to males. Investigation into the other lifestyle behaviours found that more frequent fast-food consumption and substance use were negatively associated with performance on the cognitive interference task and Math grades respectively. This study provides support for the cognitive enhancement potential of physical activity, but not for the hypothesized mediating role of brain health on academic achievement. Overall, the results speak to the importance of lifestyle behaviours in adolescent cognitive function and academic performance.

## Data Availability

Data available from corresponding author upon request.

## Acknowledgements

Thank you to Laura Xavier Fadrique who accessed and consolidated the raw Fitbit data. Thank you as well to the schools and students who participated in this study and to the teachers and administration who helped to facilitate.

